# A Prospective Evaluation of Clinical Outcomes in Acute Ischemic Stroke after Endovascular Treatment Using Transcranial Doppler (PRECISE-TCD): Study Protocol

**DOI:** 10.64898/2026.07.27.26359066

**Authors:** Song Srisilpa, Amanda Robinson, Roy Sabo, John Reavey-Cantwell, Dennis J. Rivet, Anil Roy, Shraddha Mainali, Daniel Falcao, Tarun Srivastava, Aarti Sarwal

## Abstract

Endovascular thrombectomy (EVT) improves outcomes in acute ischemic stroke caused by large vessel occlusion. Despite successful recanalization, early neurological deterioration (END) remains frequent and predicts poor outcomes. Disturbances in cerebral autoregulation may contribute to END, yet reliable bedside predictors are limited. Transcranial Doppler (TCD) provides noninvasive bedside assessment of cerebral blood flow velocities and may identify hemodynamic patterns associated with post-EVT deterioration.

PRECISE-TCD is a prospective, single-center observational study enrolling 180–300 patients undergoing EVT for anterior circulation large vessel occlusion at a tertiary academic medical center. TCD examinations will be performed as soon as feasible after EVT, daily through 72 hours, and near END events. Hemodynamic parameters including peak systolic velocity (PSV), end-diastolic velocity (EDV), mean flow velocity (MFV), and pulsatility index (PI) will be measured in bilateral MCA, ACA, PCA, carotid siphon, vertebrobasilar, and ophthalmic artery territories. The primary outcome is the association between TCD-derived parameters and END within 72 hours, defined as an increase of ≥4 points in total NIHSS score, an increase of ≥1 point in NIHSS subcategory 1a, radiologic evidence of intracranial hemorrhage within 72 hours, or any neurological change prompting emergent head CT at clinician discretion. Secondary outcomes include NIHSS at 24 hours and discharge, discharge disposition, and modified Rankin Scale (mRS) at 90 ± 10 days after hospital discharge. Joint models for longitudinal and time-to-event data will determine the association between TCD parameter trajectories and time to END. Unsupervised clustering will identify TCD-based hemodynamic phenotypes using vessel velocities, pulsatility indices, hemispheric asymmetry measures, collateral flow, and temporal trajectory patterns.

We hypothesize that abnormal post-EVT hemodynamic phenotypes will correlate with END and hemorrhagic transformation. Identifying such signatures may support development of TCD-guided, individualized blood pressure strategies to reduce secondary injury after reperfusion and inform future interventional trials. The study is registered at ClinicalTrials.gov (NCT07013396).

## Introduction

Endovascular thrombectomy (EVT) has become the standard of care for acute ischemic stroke (AIS) caused by large vessel occlusion (LVO)^1^. Extensive literature has demonstrated that EVT significantly improves functional independence at 90 days compared with medical therapy alone^2^. Despite these advances, early neurological deterioration (END) remains a frequent complication after technically successful reperfusion, with reported incidence ranging from approximately 14–35% in post-thrombectomy cohorts^3,4^. END may result from several mechanisms including infarct progression, re-occlusion, hemorrhagic transformation, symptomatic intracranial hemorrhage, or cerebral edema^3,5–7^. Current monitoring strategies rely largely on serial neurological examination and imaging with computed tomography (CT) and CT angiography. Clinical examination is often limited by baseline neurological deficits, sedation, or delirium, while imaging typically detects complications only after deterioration has already occurred. This creates an important gap for bedside physiologic monitoring capable of identifying patients at risk for secondary injury after reperfusion.

Transcranial Doppler (TCD) ultrasound provides a noninvasive bedside method for real-time assessment of cerebral blood flow velocities and cerebral hemodynamics following endovascular thrombectomy. Prior studies suggest that abnormal post-reperfusion hemodynamics identify patients at risk for secondary brain injury^8,9^. Elevated middle cerebral artery velocities, including peak systolic velocity ≥118 cm/s or an increased mean flow velocity (MFV) index relative to the contralateral hemisphere, have been associated with intracranial hemorrhage, vasogenic edema, and hyperperfusion syndromes after successful recanalization^8,9^. Other studies demonstrate that impaired cerebral autoregulation in the first 24 hours after EVT independently predicts early neurological deterioration and unfavorable functional outcomes at 90 days. Serial physiologic monitoring with TCD and other modalities shows that autoregulatory dysfunction and abnormal perfusion patterns may persist despite restoration of large-vessel patency and are associated with worse neurological recovery^10–12^. However, these investigations largely evaluate isolated physiologic signals, most commonly single-vessel MCA velocities or autoregulatory indices, without integrating comprehensive circle of Willis cerebral hemodynamics parameters accounting for collateralization across the cerebral circulation. Consequently, a coherent physiological framework to characterize post-EVT cerebral hemodynamic states remains lacking^10,11^. This limitation restricts the ability to define actionable thresholds, understand collateral flow dynamics, and translate cerebrovascular physiological monitoring into clinical decision-making after thrombectomy^12^.

Since systemic blood pressure target affects cerebral perfusion and collateralization in an impaired autoregulatory state, it has become a natural target to mitigate reperfusion injury through post-EVT blood pressure management. Disappointingly, clinical trials evaluating more intensive BP reduction have produced inconsistent results, and current AHA/ASA recommendations to maintain BP below 180/105 mmHg^13^ are largely extrapolated from thrombolysis literature rather than derived from direct physiologic monitoring. Because TCD enables bedside assessment of cerebral flow dynamics, it offers a potential strategy to tailor BP targets to patient-specific cerebral hemodynamic states rather than relying on fixed systemic thresholds. Early prospective studies have demonstrated that TCD-guided BP management is associated with lower rates of poor prognosis and higher rates of favorable functional outcomes compared with standard care^14,15^. However, these studies were conducted in Asian or European populations and primarily focus on limited hemodynamic indices within the MCA. None of these investigations evaluated a comprehensive cerebral hemodynamic profile across the Circle of Willis, incorporating collateral circulation, or performed serial assessments during the early post-reperfusion period. Consequently, physiologic thresholds that could guide individualized management after EVT remain poorly defined, particularly in U.S. stroke populations despite potential use of TCD in determining patient-specific profiles.

Key post-EVT TCD studies and the research gaps addressed by PRECISE-TCD are summarized in Table 1.

To address these gaps, we designed the PRECISE-TCD (A Prospective Evaluation of Clinical Outcomes in Acute Ischemic Stroke after Endovascular Treatment Using Transcranial Doppler) study. PRECISE-TCD is a prospective observational study evaluating post-thrombectomy cerebral hemodynamics using serial bedside TCD monitoring in patients with anterior circulation AIS undergoing EVT at a U.S. academic center. The primary objective is to determine the association between comprehensive TCD-derived parameters, including peak systolic velocity, end-diastolic velocity, mean flow velocity, and pulsatility index, and early neurological deterioration within 72 hours of EVT. The study will also examine relationships between these parameters and collateral circulation. Secondary objectives include evaluating associations between early hemodynamic patterns and clinical outcomes at hospital discharge and functional outcome at 90 ± 10 days after hospital discharge. By defining physiologic TCD thresholds and integrated cerebral hemodynamic profiles associated with END in a U.S. population, this study aims to provide foundational data for future trials of TCD-guided, individualized blood pressure management^16^ after EVT.

## Methods

### Trial Design

PRECISE-TCD is a single-center, prospective, observational study designed to identify the correlation between TCD parameters and neurological outcomes including ICH, sICH, END, and mRS scores in patients with AIS following EVT. The study will be conducted at Virginia Commonwealth University (VCU) Medical Center, Richmond, VA. The study has received IRB approval at VCU (HM20032561). The study will be conducted in accordance with the 1964 Declaration of Helsinki and its later amendments.

### Study Oversight and Dissemination

This manuscript describes the PRECISE-TCD study protocol. The trial registration record is available through ClinicalTrials.gov under NCT07013396. The study protocol and statistical analysis plan are reflected in the ClinicalTrials.gov record and institutional regulatory documents. Results from this study will be disseminated through updates to the trial registry, abstract and poster presentations at national meetings, and subsequent peer-reviewed publication. This is a prospective observational study designed to gather preliminary physiologic data. Patient and public involvement was not included in the design of this technical feasibility and observational study; patient and family input will be incorporated in future phase 2 safety and efficacy studies evaluating acceptability and implementation of TCD-guided management strategies. The study has no external sponsor or funder.

### Inclusion and Exclusion Criteria

Eligible patients must meet all of the following: (1) Age ≥18 years; (2) Anterior circulation LVO, including ACA, MCA, or ICA stroke, treated with EVT, including tandem occlusions; (3) Ability to detect an adequate acoustic window via TCD.

Patients will be excluded if they: (1) Have inadequate acoustic windows defined as lack of bilateral MCA signal at standard depths; (2) Are pregnant; (3) Are incarcerated; or (4) Have already been transitioned to a comfort-care pathway.

### Informed Consent

Informed consent will be obtained electronically via the REDCap e-Consent platform approved by the IRB, in person in the Emergency Department or Neurocritical Care Unit prior to any research procedures. Participants will be given ample time to decide with no imposed time limit. If a participant lacks decision-making capacity, consent will be obtained from a legally authorized representative (LAR). Reconsent will be obtained from the participant if capacity is later regained. Participants may withdraw at any time before research-related activities begin. No biological specimens are collected as part of this study, and no ancillary biological specimen consent is applicable.

### Recruitment and Retention

Eligible participants will be identified through collaboration with the neurocritical care, stroke neurology, and endovascular teams involved in the care of patients with acute ischemic stroke undergoing EVT. Dedicated research coordinator support will be used to screen potentially eligible patients, coordinate consent, and facilitate completion of study-related TCD assessments. Participant retention and follow-up will be promoted through regular engagement with frontline providers, research coordinators, participants, and families to support completion of in-hospital assessments and the outcome follow-up conducted 90 ± 10 days after hospital discharge.

### TCD Measurements and Data Collection

TCD examinations will be performed as soon as feasible after EVT, daily through 72 hours, and, when feasible, near any reported neurological deterioration or clinically indicated neuroimaging within 72 hours of EVT. The schedule of enrolment, study procedures, and assessments is shown in Fig 1.

**Fig 1.**
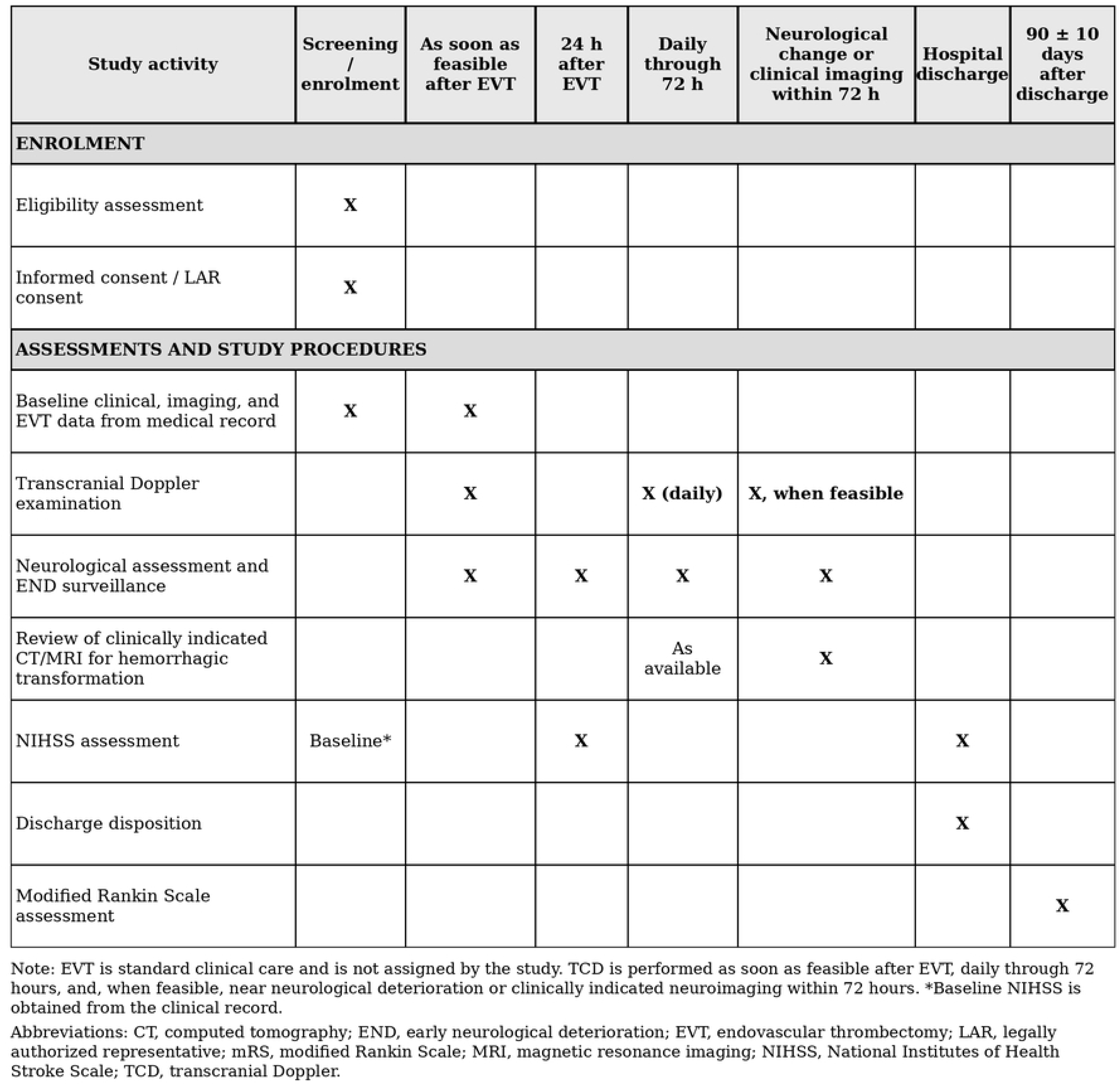
SPIRIT schedule of enrolment, study procedures, and assessments for PRECISE-TCD. EVT is standard clinical care and is not assigned by the study. TCD examinations are performed as soon as feasible after EVT, daily through 72 hours, and, when feasible, near neurological deterioration or clinically indicated neuroimaging within 72 hours. The 90-day mRS follow-up is conducted 90 ± 10 days after hospital discharge. *Baseline NIHSS is obtained from the clinical record. Abbreviations: CT, computed tomography; END, early neurological deterioration; EVT, endovascular thrombectomy; LAR, legally authorized representative; mRS, modified Rankin Scale; MRI, magnetic resonance imaging; NIHSS, National Institutes of Health Stroke Scale; SPIRIT, Standard Protocol Items: Recommendations for Interventional Trials; TCD, transcranial Doppler.

All studies will be conducted by a trained sonographer using low-frequency MHz TCD probes. Vessels assessed will include the bilateral MCA, ACA, PCA, carotid siphon (Siph), vertebrobasilar arteries and ophthalmic artery (OA). TCD parameters collected will include: PSV, EDV, MFV, and PI. The sampling volume will be 5–10 mm. Monitoring depth on transtemporal windows will be adjusted per vessel: distal MCA 40–50 mm, proximal MCA 50–60 mm, ACA 70–75 mm, PCA 62–70 mm; transorbital: Siph 60– 64 mm, OA 50–60 mm. TAMMV (time-averaged mean flow velocity) and PI will be automatically derived from preset values. All vessel segments insonated will be recorded. Any evidence of collateralization as evidenced by ipsilateral reversed ACA, ipsilateral hyperemic PCA, contralateral hyperemic ACA or ipsilateral reversed ophthalmic artery will be noted. Table 2 summarizes the key hemodynamic domains captured by TCD: flow magnitude, downstream resistance, hemispheric symmetry, and collateral circulation, providing a framework to interpret physiologic changes after reperfusion therapy. The conceptual framework linking serial TCD measurements, hemodynamic domains, and clinical outcomes is shown in Fig 2.

**Fig 2.**
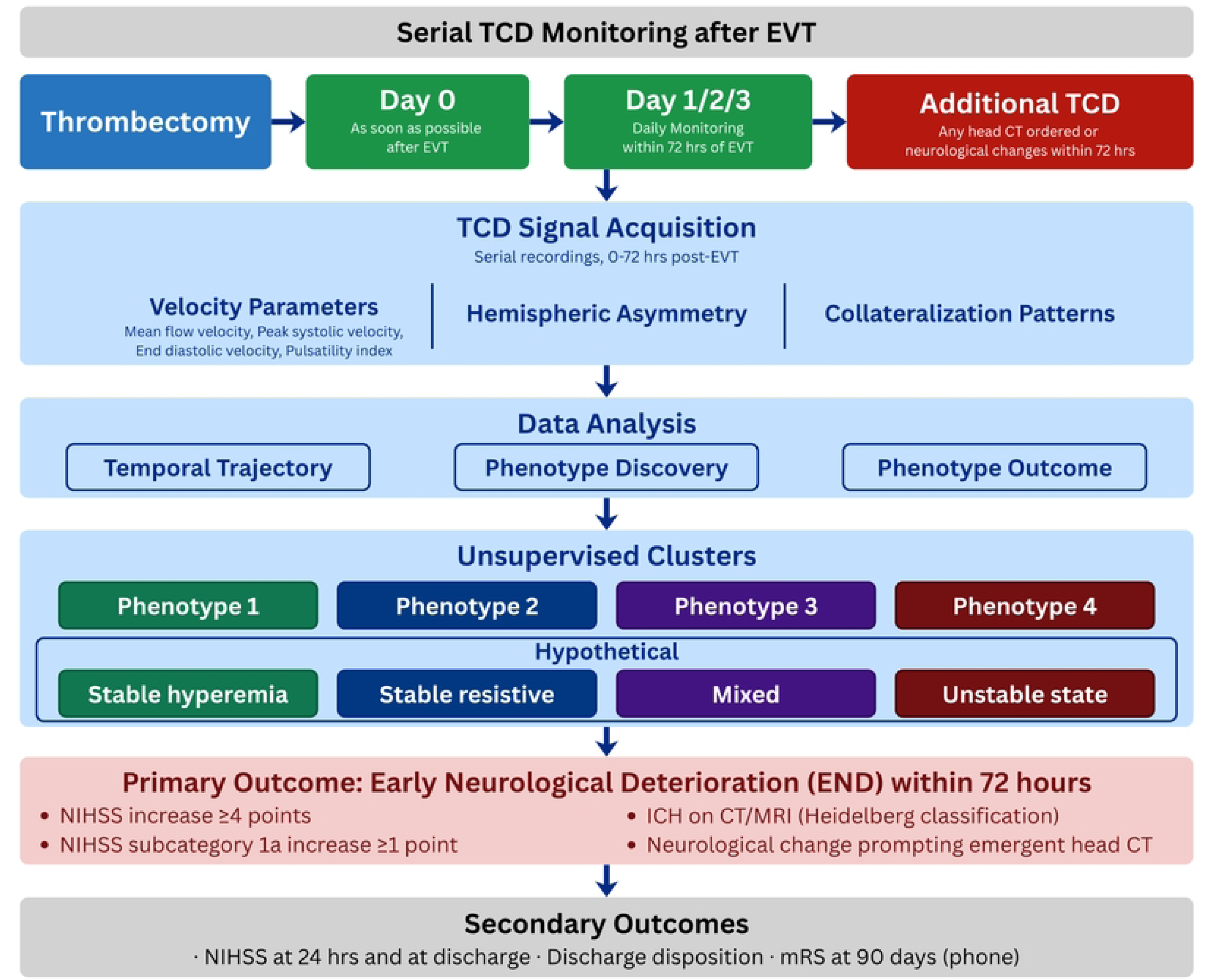
Conceptual framework for transcranial Doppler monitoring after endovascular thrombectomy. Serial transcranial Doppler (TCD) monitoring is performed as soon as feasible after endovascular thrombectomy (EVT), daily through 72 hours, and, when feasible, near neurological deterioration or clinically indicated neuroimaging within 72 hours. Extracted parameters involve three hemodynamic domains: velocity metrics (PSV, EDV, MFV, and PI), hemispheric asymmetry, and collateralization patterns. Together, these parameters may characterize different cerebral perfusion states, including stable or unstable hypoperfusion or hyperperfusion states in patients with varying degrees of collateralization. Integrating these signals and adjusting for patient factors may help detect ischemic deterioration and hemorrhagic transformation, guide blood pressure management, and identify patients at risk for early neurological deterioration. Abbreviations: CT, computed tomography; EDV, end-diastolic velocity; END, early neurological deterioration; EVT, endovascular thrombectomy; MFV, mean flow velocity; PI, pulsatility index; PSV, peak systolic velocity; TCD, transcranial Doppler.

All studies will adhere to ALARA principles and AIUM safety guidelines^17,18^. Collateralization status will be assessed as an indirect indicator of arterial occlusion or severe stenosis^19,20^ in M1 MCA, identified by: (1) reversal of flow in the ipsilateral ACA; (2) high-velocity pattern in the ipsilateral ACA or PCA (ACA/PCA MFV > MCA MFV, with ACA/PCA stenosis ruled out from prior CTA/DSA); and (3) reversal of the ipsilateral ophthalmic artery^14,15^. The monitoring schedule was designed to capture the period when reperfusion-related hemodynamic instability is most likely to occur after thrombectomy. Prior studies demonstrate that dynamic cerebral autoregulation abnormalities can be detected within the first 24 hours after EVT and are associated with neurological and functional outcomes^21^, while autoregulatory dysfunction after ischemic stroke may persist for several days during the early recovery phase^22^. In addition, hyperperfusion phenomena after cerebral revascularization procedures most commonly develop within the first 24–72 hours and can be detected with serial Doppler monitoring^9^. Accordingly, TCD monitoring as soon as feasible after EVT, with daily assessments through 72 hours, was selected to capture this critical window of evolving cerebral hemodynamics.

To minimize operator-dependent variability, all serial TCD examinations will be performed using a standardized acquisition protocol with predefined insonation depths, probe orientation, and velocity measurements across target vessels. Sonographers and investigators performing the studies will undergo structured training and calibration prior to study initiation. Periodic quality assurance reviews will be conducted, including blinded re-measurement of a subset of recordings to assess intra-rater reliability. All waveforms will be stored digitally and reviewed when needed to ensure adherence to protocol and consistency of velocity measurements across time points.

### Harms and Safety Monitoring

TCD is a noninvasive, radiation-free bedside ultrasound modality with an established safety profile. This observational study does not alter standard clinical care or assign participants to an intervention. Therefore, no study-related harms are anticipated beyond the minimal risks associated with bedside ultrasound assessment. Any unexpected study-related concerns will be reviewed by the study team and managed according to institutional and IRB requirements.

## Outcomes

### Primary outcome

END is defined as: (1) an increase in total NIHSS score of ≥4 points; (2) an increase in NIHSS subcategory 1a score of ≥1 point; (3) incidence of ICH within 72 hours of EVT on CT or MRI classified through Heidelberg Classification for hemorrhagic conversion in ischemic stroke, or (4) any change in examination inciting need for an emergent head CT based on clinician discretion within 72 hours post-thrombectomy.

### Secondary outcome

(1) Post-EVT NIHSS score at 24 hours and at discharge. (2) Discharge disposition (home, acute rehabilitation, long-term acute care facility, skilled nursing facility, or hospice/palliative care/death). (3) Functional outcome assessed by phone-based mRS at 90 ± 10 days after hospital discharge (good outcome: mRS 0–2; poor outcome: mRS 3–6).

### Sample Size Justification

The sample size for PRECISE-TCD was determined based on the expected incidence of early neurological deterioration after EVT and the need to detect clinically meaningful associations between TCD-derived hemodynamic profiles and END. Prior studies report END rates of approximately 20–30%^3,4^. Assuming an event rate of 25%, enrollment of approximately 180 patients would yield about 45 END events, providing adequate power (α=0.05, 80% power) to detect moderate associations (odds ratio ∼2.0) between abnormal TCD profiles and END while allowing parsimonious multivariable modeling consistent with recommended event-to-predictor ratios. Specifically, prioritizing 2 to 4 clinically primary covariates to maintain an event-to-variable ratio consistent with established statistical recommendations (10-20 events per degree of freedom)^23^. END incidence of less than 10% following thrombectomy will need enrollment of approximately 300 patients to yield 24–30 END events.

An interim analysis will be performed after approximately 50% of the planned cohort is enrolled to verify the observed incidence of early neurological deterioration and assess completeness and quality of serial TCD data acquisition. This analysis will be descriptive only and will not include formal hypothesis testing; if the observed END incidence falls substantially below the anticipated 10–25% range, adjustment of the final sample size may be considered to ensure adequate numbers of outcome events for the planned exploratory analyses.

This sample size also supports exploratory receiver operating characteristic (ROC) analyses to identify candidate cutoff values for multiparameter TCD metrics, including PSV, EDV, MFV, PI, hemispheric asymmetry, and collateral flow patterns—measured during the first 72 hours after EVT. Although not designed for definitive model validation, this cohort is expected to provide sufficiently robust physiologic signal detection to inform future multicenter trials evaluating TCD-guided individualized blood pressure management after thrombectomy.

## Statistical Analysis

Continuous variables will be summarized as mean ± standard deviation or median with interquartile range, and categorical variables as counts and percentages. Baseline characteristics will be compared between patients with and without early neurological deterioration using Student’s t-test or Mann–Whitney U test for continuous variables and chi-square or Fisher’s exact test for categorical variables.

TCD-derived hemodynamic parameters (PSV, EDV, MFV, and PI) will be analyzed at both the vessel level and the patient level. Measurements from bilateral MCA, ACA, PCA, carotid siphon, vertebrobasilar arteries and bilateral ophthalmic arteries will be recorded for each time point. Derived variables will include ipsilateral-to-contralateral velocity ratios, inter-vessel velocity relationships (e.g., ACA/MCA and PCA/MCA ratios), and asymmetry indices between hemispheres. The maximum MCA velocity on both the ipsilateral and contralateral sides will serve as the consistent patient-level measure for serial analysis. Collateralization patterns will be defined based on established TCD criteria including reversal of the ipsilateral ACA or ophthalmic artery and high-flow patterns in contralateral ACA or ipsilateral PCA relative to MCA flow velocities.

We will use joint models for longitudinal and time-to-event data to explore the association between changes in TCD parameters and time to END in hours. We will use Bayesian methods to estimate the hazard ratio from a semi-parametric proportional hazard regression model for a shared patient-level random slope obtained from a linear random effects model predicting individual TCD parameters. With TCD parameters being collected daily over three days, we will be limited to looking at linear changes, corresponding to a constant rate over time. This framework will allow us to model the trajectory of TCD parameters even for those individuals with fewer than three measurements as well as to account for incomplete measurements for early events and death censoring. We will consider modelling additional covariates including age, baseline NIHSS, occlusion location, and other clinical factors with the goal of avoiding an overfit model.

To identify integrated cerebral hemodynamic phenotypes, unsupervised clustering methods will be applied to standardized TCD features including vessel velocities, pulsatility indices, hemispheric asymmetry measures, collateral flow indicators, and temporal trajectory patterns over the monitoring period. Because post-EVT cerebral physiology reflects interacting domains of flow magnitude, vascular resistance, hemispheric balance, and collateral recruitment, traditional single-parameter analyses may not capture the full spectrum of physiologic states. Data-driven clustering therefore enables discovery of latent hemodynamic profiles without prespecified thresholds. We will use finite mixture modelling to determine phenotype weights for each subject for each of *g* phenotype groups. The number of phenotype groups “g” can be determined empirically by selecting the number of groups that minimizes the Bayesian information criterion (BIC)^24^. To assess model discrimination, or the ability of our probability-based model to identify unique groups of patient phenotypes, we will estimate average posterior probabilities (APP) and odds of correct classification (OCC); models with APPs greater than 0.7 are generally considered admissible^25,26^. We hypothesize that this unsupervised analysis will complement observed hemodynamic profiles consistent with patterns observed in the post-EVT physiology literature. Current presumed phenotypes based on limited literature include stable hyperaemic state characterized by elevated flow velocities without increased resistance; a stable resistive state reflecting high downstream vascular resistance; a mixed phenotype combining features of both hyperemia and resistance; and an unstable State defined by dynamic transitions between hemodynamic profiles across serial assessments. Intersection of these phenotypes with collateralization status has not been studied. The phenotypes resulting from unsupervised clustering that integrate more holistic cerebrovascular profile will be evaluated for associations with early neurological deterioration, haemorrhagic transformation, and clinical outcomes. Secondary analyses will evaluate relationships between early TCD-derived hemodynamic patterns and functional outcomes including NIHSS at 24 hours and discharge, discharge disposition, and modified Rankin Scale score at 90 ± 10 days after hospital discharge using logistic or ordinal regression models as appropriate. Additional analyses will examine interactions between TCD-derived phenotypes, and collateralization status to determine whether this impairment modifies the relationship between cerebral hemodynamic and clinical outcomes. Because serial physiological monitoring may yield intermittent missing observations, repeated TCD measures will be analyzed using mixed-effects models that allow inclusion of patients with incomplete time points. Missing covariates will be addressed with multiple imputation if needed, and sensitivity analyses will compare results with complete-case models. Patients without any post-EVT TCD data will be excluded from longitudinal analyses.

Disclosure of Use of AI: In accordance with PLOS ONE policy and ICMJE recommendations, the authors disclose that AI-assisted technologies (Copilot, Microsoft; ChatGPT, OpenAI; Claude, Anthropic) were used during manuscript preparation for editorial purposes only, including copy-editing and language polishing of author-drafted text, structural and visual formatting of the manuscript file and tables, and consistency checking of in-text citations against the reference list. No AI tool was used to generate scientific content, design the study described, conduct any analysis, or draft conclusions. All scientific content, data, and conclusions are those of the human authors, who take full responsibility for the accuracy, integrity, and originality of the work.

## Discussion

PRECISE-TCD addresses a key gap created by the rapidly emerging literature on post-EVT cerebral hemodynamics. Prior studies have suggested that abnormal TCD flow patterns may identify patients at risk for reperfusion injury, but these investigations have largely focused on single parameters, limited vascular territories, or non-U.S. populations, and have not established physiologic thresholds that could support actionable bedside monitoring^8,9^. As a result, the field lacks an integrated framework for translating TCD observations into individualized management strategies. PRECISE-TCD is designed to address this gap by prospectively characterizing multiparameter cerebral hemodynamic profiles after EVT in a contemporary U.S. stroke population. The study systematically evaluates PSV, MFV, EDV, and PI across the anterior and posterior circulation, including bilateral MCA, ACA, PCA, ophthalmic, carotid siphons and vertebrobasilar arteries, while incorporating collateral flow patterns and serial assessments across the full 72-hour post-reperfusion window. By integrating velocity patterns, pulsatility metrics, and collateral signatures, the study aims to define physiological TCD profiles associated with early neurological deterioration and functional outcomes. By establishing these hemodynamic patterns and candidate thresholds, PRECISE-TCD provides the next critical step toward translating noninvasive cerebrovascular monitoring into individualized patient care. The resulting data will help define how bedside TCD can be used to stratify post-EVT patients according to their cerebral hemodynamic state and will generate the physiological framework necessary to design future trials of TCD-guided, patient-specific blood pressure management strategies.

The existing post-EVT TCD literature is promising but remains limited in scope. He et al. (2020) demonstrated in a Chinese cohort that elevated middle cerebral artery velocities, specifically PSV ≥118 cm/s and an MFV index ≥1.12, were independently associated with intracranial hemorrhage and vasogenic edema following successful recanalization^8^. Kneihsl et al. (2018) similarly reported in an Austrian cohort that an MFV index ≥1.25 predicted cerebral hyperperfusion syndrome after thrombectomy, a condition that may precede hemorrhagic transformation^9^. Prospective interventional work by Chen et al. (2020) and Tao et al. (2021) further demonstrated that TCD-guided blood pressure management in Chinese patients resulted in significantly improved functional outcomes at 90 days compared with standard care^14,15^. Additional studies have expanded the physiological insights obtained from TCD monitoring after EVT. Tian et al. (2020) showed that impaired dynamic cerebral autoregulation measured using TCD shortly after EVT independently predicted unfavorable functional outcomes at 90 days, highlighting persistent hemodynamic instability despite successful recanalization^21^. Subsequent work has reinforced this observation, demonstrating that abnormal autoregulatory metrics measured serially with TCD are associated with worse neurological recovery. Studies incorporating multimodal physiological monitoring have also explored the integration of TCD with other techniques. For example, Qi et al. (2024) evaluated combined TCD and quantitative EEG (qEEG) monitoring as an early marker of futile recanalization, identifying physiologic signatures associated with poor neurological recovery despite technical reperfusion success^27^. Early post-EVT studies have demonstrated that persistent collateral recruitment or flow diversion detected on TCD, such as reversed anterior cerebral artery or ophthalmic artery flow, may be associated with early neurological deterioration and unfavorable outcomes, suggesting that collateral-dependent flow states may persist even after large-vessel recanalization. These observations support the concept that cerebral hemodynamics after EVT reflect a complex interaction between reperfusion, autoregulatory function, and collateral circulation^19,20^. Taken together, these studies establish a consistent signal that post-EVT TCD has both prognostic and potentially interventional value. However, several key limitations remain. Most studies have been conducted in Asian or Central European populations, many have focused on single hemodynamic parameters or the MCA territory alone, and few have systematically incorporated collateral circulation into outcome models. Furthermore, none have generated validated multiparameter TCD thresholds applicable to contemporary U.S. stroke populations. These gaps in physiological characterization and population-specific validation are precisely the areas that the PRECISE-TCD study is designed to address.

PRECISE-TCD addresses several gaps created by these studies. Most prior studies evaluate isolated physiological signals, typically single-vessel MCA velocities, without assessing the broader cerebral circulation. This is an important limitation because EVT alters flow redistribution across the entire Circle of Willis, the principal intracranial collateral network linking anterior and posterior circulations^19,20^. Hemodynamic responses in adjacent territories, including the carotid siphon and posterior circulation, may therefore reflect collateral recruitment or compensatory flow shifts when anterior circulation perfusion changes^10,21^. Transcranial Doppler can detect these collateral pathways by identifying velocity changes or flow direction reversal across communicating arteries and posterior circulation vessels^19,20^. Such global flow patterns may precede clinical deterioration and therefore provide additional predictive value for early neurological deterioration beyond MCA velocities alone. In addition, patients may show stable or fluctuating patterns of perfusion abnormalities or collateral patterns. Stability of perfusion patterns has been shown to be consequential in post-cardiac arrest TCD literature and has prognostic significance^21^. Consequently, an integrated multi-territory TCD framework may better characterize post-EVT cerebral hemodynamic states and improve risk stratification after reperfusion.

This study has several limitations. First, as a single-center observational study, selection bias and center-specific practice patterns may limit generalizability. However, standardized EVT protocols and uniform neurocritical care management at the study site allow consistent physiological monitoring and reduce variability in post-reperfusion care. Second, serial TCD monitoring is operator-dependent. This was mitigated through a standardized acquisition protocol, structured training of operators, and periodic waveform review to ensure measurement consistency. Third, intermittent missing physiological measurements may occur during serial monitoring. Longitudinal mixed-effects modeling was prespecified to accommodate incomplete repeated measures without excluding patients with partial data. Finally, although the study may be underpowered to detect very rare complications, the sample size was designed to evaluate clinically meaningful associations between post-EVT hemodynamic patterns and early neurological deterioration. We also do not directly evaluate autoregulatory status, despite the potential promise of this technology and emerging literature of the potential role of determining autoregulatory status in these patients. Direct autoregulation assessment using TCD monitoring with carotid compression is not widely accepted in routine clinical practice, and multimodal autoregulation monitoring tools remain limited in availability requiring high resources. Our goal was to develop a scalable serial TCD-based algorithm using parameters that can be derived directly from a complete bedside TCD examination which are more feasible to perform in routine neurocritical care practice. This pragmatic approach prioritizes measures that are feasible, reproducible, and implementable across real-world clinical settings. These limitations should therefore be interpreted in the context of a physiologically detailed monitoring framework that provides granular insight into post-reperfusion cerebral hemodynamics.

Despite these limitations, PRECISE-TCD represents an important step toward addressing a critical evidence gap in post-EVT monitoring. By prospectively defining multiparameter TCD thresholds and integrating collateral flow patterns into outcome analyses, the study aims to characterize physiological cerebral hemodynamic profiles after reperfusion. These data are necessary to define clinically relevant phenotypes, that may suggest intervenable cerebrovascular profiles like stable hyperemia, or stable resistive flows with prespecified collateralization states and will provide the foundation for future trials of TCD-guided, individualized blood pressure management strategies aimed at reducing early neurological deterioration and hemorrhagic complications after EVT.

## Conclusions

Existing evidence suggests that post-EVT TCD markers including abnormal velocity thresholds, impaired autoregulation, and persistent collateral flow patterns are associated with reperfusion injury and poor outcomes; however, most studies evaluate limited MCA-based parameters, and TCD-guided blood pressure strategies have primarily been tested in Asian cohorts, leaving a need for comprehensive multiparameter assessment across the Circle of Willis in broader populations. PRECISE-TCD is a prospective observational study evaluating the relationship between serial TCD-derived hemodynamic parameters and early neurological deterioration in patients with anterior circulation LVO stroke after EVT at a U.S. academic center. By defining physiological TCD thresholds in a population underrepresented in prior studies, this work aims to provide the empirical basis for future trials of TCD-guided, individualized blood pressure management to reduce neurological deterioration and hemorrhagic complications after thrombectomy.

## Declarations

Ethics Approval and Consent to Participate: The original study protocol, informed consent documents, case report forms, and supporting materials were reviewed and approved by the VCU IRB under HM20032561. The study will be conducted in accordance with the ethical standards of the 1964 Declaration of Helsinki and its later amendments. Informed consent will be obtained from all participants or their legally authorized representatives. Important protocol modifications will be submitted for IRB review and approval before implementation when required, and relevant updates will also be reflected in the ClinicalTrials.gov record.

## Consent for Publication

Not applicable.

## Availability of Data and Materials

No datasets have been generated or analyzed because recruitment has not yet begun. De-identified data generated during the study may be made available from the corresponding author upon reasonable request, subject to institutional and ethical requirements.

## Author Contributions

Song Srisilpa (SS): Data analysis, project coordination, data collection, regulatory management, data management, data entry, data coding, and contribution to study design.

Amanda Robinson (ARo): Study design, statistical analysis planning, and development of the analytic framework.

Roy Sabo (RS): Study design, statistical analysis planning, and development of the analytic framework.

John Reavey-Cantwell (JRC): Participant identification, study design, and provision of clinical services.

Dennis Rivet (DR): Participant identification, study design, and provision of clinical services.

Anil Roy (AR): Participant identification, study design, and provision of clinical services.

Shraddha Mainali (SM): Participant consent, participant identification, study design, and provision of clinical services.

Daniel Falcao (DF): Participant consent, participant identification, study design, and provision of clinical services.

Tarun Srivastava (TS): Manuscript preparation, data management, participant consent, participant identification, study coordination, and corresponding author.

Aarti Sarwal (AS): Study design, data analysis, participant consent, and overall study supervision.

All authors meet the ICMJE criteria for authorship. The final manuscript has been approved by all authors.

Reporting Checklist: A SPIRIT checklist is included as a supplementary document given the study protocol nature of this submission.

## Competing Interests

Aarti Sarwal: Butterfly Network, Inc. and Image Monitoring Inc. have provided loaned ultrasound devices to support investigator-initiated research conducted by Dr. Sarwal. Dr. Sarwal serves as a consultant and member of the Clinical Events Committee for the REINVIGORATE trial (REINVIGORATE Trial; NCT05998018), sponsored by Stimdia Medical Inc. She also serves as a consultant and member of the Data Safety Monitoring Board for the phase II MR-301 trial (NCT06253923), sponsored by Shinkei Therapeutics Inc.

Shraddha Mainali: Image Monitoring Inc has provided a research loan of devices to support an investigator-initiated trial.

The remainder of the authors declare that they have no conflicts of interest.

## List of Abbreviations

ACA: Anterior cerebral artery
AIS: Acute ischemic stroke
ALARA: As low as reasonably achievable
AIUM: American Institute of Ultrasound in Medicine
BP: Blood pressure
CAR: Cerebral autoregulation
CBF: Cerebral blood flow
CI: Confidence interval
CT: Computed tomography
dCAR: Dynamic cerebral autoregulation
EDV: End-diastolic velocity
END: Early neurological deterioration
EVT: Endovascular thrombectomy
ICA: Internal carotid artery
ICH: Intracranial hemorrhage
IQR: Interquartile range
IRB: Institutional Review Board
LAR: Legally authorized representative
LVO: Large vessel occlusion
MCA: Middle cerebral artery
MFV: Mean flow velocity
MRI: Magnetic resonance imaging
mRS: Modified Rankin Scale
NIHSS: National Institutes of Health Stroke Scale
OA: Ophthalmic artery
OR: Odds ratio
PCA: Posterior cerebral artery
PI: Pulsatility index
PSV: Peak systolic velocity
qEEG: Quantitative electroencephalography
ROC: Receiver operating characteristic
SD: Standard deviation
sICH: Symptomatic intracranial hemorrhage
Siph: Carotid siphon
TAMMV: Time-averaged mean flow velocity
TCD: Transcranial Doppler
TICI: Thrombolysis in cerebral infarction
VCU: Virginia Commonwealth University

## Data Availability

No datasets were generated or analysed during the current study. All relevant data from this study will be made available upon study completion.

## Acknowledgments

Not applicable.

## Supporting Information

S1 Checklist. Completed SPIRIT 2025 checklist.

S2 Protocol. PRECISE-TCD study protocol.

